# Cardiac rhythm development: A wearable device index of risk for physical and mental illness in adolescence

**DOI:** 10.64898/2026.06.12.26355405

**Authors:** Eugenia Giampetruzzi, Katharina Kircanski, Daniel S. Pine, Ian H. Gotlib

## Abstract

**Objective:** The autonomic nervous system, which regulates cardiac rhythm, undergoes pronounced maturation across adolescence. How cardiac rhythm develops over this period, however, and whether individual differences in its development forecast mental and physical illness, remain open questions. We used three waves of Fitbit data from the Adolescent Brain Cognitive Development (ABCD) Study to characterize the developmental trajectory of the cardiac rhythm and to test whether variation in that trajectory predicts onset of psychopathology and cardiometabolic disease.

**Methods:** 8,301 adolescents contributed 242,811 valid Fitbit wear days across Waves 2 (Mage=12), 4 (Mage=14), and 6 (Mage=16). Cosinor mixed-effects models yielded three rhythm parameters per session: mesor (24-hour mean), amplitude (diurnal swing), and acrophase (peak timing). We first characterized age- and sex-specific trajectories, cross-wave stability, and factors shaping the rhythm. We then used parallel-process latent growth models to test whether within-person changes in rhythm tracked symptom trajectories, and hierarchical logistic models to test whether rhythm parameters predicted the first clinical onset of psychopathology and of obesity and hypertension.

**Results:** The cardiac rhythm changed substantially across adolescence: mesor decreased, amplitude flattened, and acrophase shifted later. Within-person change in the rhythm tracked change in blood pressure, BMI, and trajectories of depression and ADHD symptoms. Higher mesor predicted incident onset of all five outcomes controlling for demographics, baseline symptoms, and behavior (ORs 1.36-1.54); amplitude, acrophase, and rhythm instability conferred additional risk.

**Conclusions:** The 24-hour cardiac rhythm is a passively measurable substrate of adolescent autonomic development that indexes transdiagnostic risk for psychiatric and cardiometabolic illness.

## Introduction

The autonomic nervous system (ANS) undergoes pronounced maturation across adolescence (Dollar et al., 2020; Gibbons et al., 2019). Its regulation of the heart gives cardiac activity a marked 24-hour rhythm: heart rate rises during activity, peaks at a consistent time each day, and falls during sleep (Hadtstein et al., 2004). This rhythm is reshaped across adolescence as cardiac autonomic control matures. While this change is normative, its pace and form vary considerably across individuals. The factors that give rise to this variability, and its possible effects on health, are not well understood.

The broad course of cardiac autonomic maturation has been described in two large-scale studies. First, in the most comprehensive laboratory-based investigation to date, Harteveld and colleagues (2021) characterized maturational trajectories of cardiac activity across 4,820 healthy individuals from infancy to early adulthood. Second, in a meta-analysis of 45 studies (3,886 youth) using 24-hour Holter monitoring, Joyce et al. (2025) found that heart rate decreased consistently with age from infancy to early adulthood. Importantly, both studies also reported substantial interindividual variability at all ages. This variability is unlikely to be random: cardiac autonomic regulation is shaped by lifestyle behaviors such as physical activity (Tornberg et al., 2019) and sleep (Morales-Ghinaglia et al., 2025), as well as by the broader socioeconomic and demographic contexts in which adolescents are raised (Chaix et al., 2010). Thus, the developmental course of ANS functioning reflects a combination of intrinsic autonomic maturation and the behavioral and contextual forces that influence it.

It is important that we characterize this development and trajectory given that psychiatric and cardiometabolic difficulties first emerge in adolescence, share cardiac ANS imbalance as a developmental risk factor, and frequently co-occur into adulthood (Galler et al., 2024; Kwapong et al., 2023; Muha et al., 2026). Elevated resting heart rate and reduced heart rate variability have been associated with a range of psychiatric conditions (Alvares et al., 2016), obesity (Freitas Júnior et al., 2012), and hypertension (Liu et al., 2021), implicating ANS imbalance as a transdiagnostic risk factor. We do not yet know, however, whether individual differences in the development of the rhythm across adolescence forecast the emergence of these difficulties, in part because of how researchers have measured cardiac activity. Brief laboratory recordings, typically under 10 minutes, capture only a static snapshot of autonomic function, and 24-hour clinical Holter parameters are reported as discrete reference values; neither characterizes the shape of the diurnal rhythm itself. Cosinor modeling parameterizes that shape, yielding the rhythm-adjusted 24-hour mean (mesor), the diurnal swing (amplitude), and the clock time of the peak (acrophase), features that index physiological flexibility and cumulative regulatory burden in ways that scalar summaries cannot (Hadtstein et al., 2004). To date, cosinor parameterization of cardiac rhythm has been applied at scale longitudinally in adults (N=92,000; Quer et al., 2020) but only cross-sectionally in youth (Hadtstein et al., 2004 [N=983]; Sigrist et al., 2024 [N=30]). Researchers have not yet characterized how the cosinor-parameterized 24-hour rhythm develops normatively across adolescence and whether individual differences in that development have prognostic significance for mental and physical health.

Consumer wearables can address these questions by enabling continuous, passive characterization of the 24-hour cardiac rhythm in everyday settings. The Adolescent Brain Cognitive Development (ABCD) Study began collecting Fitbit data at the two-year follow-up, capturing roughly 21 days of cardiac data per participant across waves in a national sample of nearly 12,000 adolescents (Bagot et al., 2018; Karcher & Barch, 2021). Thus far, however, research using these data has relied on device-computed scalar summaries at only one wave (e.g., Damme et al., 2024). In the present study we used three waves of ABCD Fitbit data to characterize how the 24-hour cardiac rhythm develops across adolescence and to test its prognostic value. We hypothesized, first, that the cardiac rhythm will mature systematically with age, with the 24-hour mean declining across adolescence consistent with the developmental decrease in heart rate documented in prior work (Joyce et al., 2025), while remaining sufficiently stable across waves to serve as a trait-like individual marker of vulnerability to disorder. Second, we hypothesized that a higher, flatter rhythm in early adolescence will index transdiagnostic risk, forecasting both the subsequent trajectory and the first clinical onset of psychopathology and cardiometabolic disease two to four years later, beyond the variance explained by demographic and behavioral variables.

## Methods

### Participants

*Overview*. Data were drawn from the Adolescent Brain Cognitive Development (ABCD) Study, in which children were recruited at ages 9-10 from 21 U.S. sites (Karcher & Barch, 2020). At the two-year follow-up, a subset of adolescents entered a Novel Technologies sub-study and were issued a Fitbit Charge HR2 to wear continuously, including during sleep, for 21 days; collection continued at the four- and six-year follow-ups (Bagot et al., 2018). The full sample was composed of 8,301 adolescents contributing 13,552 quality-criteria-meeting Fitbit sessions across Waves 2 (N=7,230), 4 (N=4,306), and 6 (N=2,016) yielding 371 million minute-level heart rate readings. Developmental analyses used the full sample. Co-development analyses used the Wave-2 cosinor cohort (N=7,230), with full-information maximum likelihood for missing follow-up data. Onset analyses compared healthy controls (N=1,188 below clinical threshold on all outcomes at every wave) with 1,360 distinct adolescents who developed an incident clinical elevation at Waves 4 or 6. All ABCD procedures were IRB-approved; parents provided informed consent and youth provided assent.

#### Demographics

Demographic data were obtained from caregivers at baseline: youth sex assigned at birth, race and ethnicity (White, Hispanic, Black, Asian, Other), household income (three categories), and the highest education attained by either caregiver (five categories). ABCD recruitment site (21 sites) was used as a clustering variable in all multilevel analyses.

### Wearable device

#### Protocol

The Fitbit Charge HR2 records cardiac and activity data via photoplethysmography at a one-second sampling rate (Bagot et al., 2018; Damme et al., 2024). ABCD releases heart rate as minute-level averages aggregated from this stream, along with daily step counts and sleep duration. We used minute-level heart rate to model the cardiac rhythm and daily steps and total sleep duration as covariates.

#### Quality criteria

Sessions were retained if they contained at least three valid wear days, defined as days with at least 600 minutes of non-zero heart rate data distributed across all four six-hour clock-time quadrants (00:00-06:00, 06:00-12:00, 12:00-18:00, 18:00-24:00) to ensure sufficient temporal coverage of the 24-hour cycle.

### Outcome Variables

#### Psychopathology

Psychopathology was assessed at each wave using two measures: the Child Behavior Checklist (CBCL; Achenbach & Rescorla, 2001) and the Kiddie Schedule for Affective Disorders and Schizophrenia for DSM-5 (KSADS-COMP; Townsend et al., 2020). The CBCL is a 113-item parent-report measure that yields scales scored along several dimensions, and the KSADS-COMP is a parent-reported assessment of DSM diagnoses. We examined the three categories that are assessed by both instruments: depression, anxiety, and externalizing problems. Due to differential response rates, we used the CBCL as the primary measure and the KSADS-COMP as an exploratory convergence check; we also analyzed the child-reported KSADS-COMP to compare findings obtained through parent report. The CBCL yields DSM-oriented scales with age- and sex-normed T-scores; a T-score ≥ 65 (≥93rd percentile) defines the borderline-clinical range (Achenbach & Rescorla, 2001), which was used as the clinical threshold for the binary-onset analyses. We used the Depressive Problems and Anxiety Problems scales for both the continuous and binary-onset analyses. For the onset analyses, externalizing was a combined category (scored as a clinical-range elevation on the Attention-Deficit/Hyperactivity [ADHD], Oppositional Defiant [ODD], or Conduct Problems [CDD] scales); for the trajectory analyses, these three externalizing problems scales were examined separately. The KSADS-COMP is a validated, computerized self-administered version of the KSADS, on which each of the three diagnoses assessed in this study is coded as present (1) or absent (0)

#### Cardiometabolic disease

We assessed two primary cardiometabolic outcomes: obesity, indexed by body mass index (BMI), and hypertension, indexed by blood pressure (BP). Consistent with prior work with ABCD (Al-Shoaibi et al., 2024; Tomasi & Volkow, 2024), obesity was defined as BMI at or above the 85th percentile for age and sex, and hypertension was defined as systolic and/or diastolic blood pressure ≥95th percentile for children younger than 13, or ≥130/80 mm Hg for children 13 and older, both per CDC growth charts and guidelines.

## Data Analysis

Analyses were conducted in Python 3.12 and R 4.5.2 (cosinor mixed-effects models, lme4; latent growth and parallel-process models, lavaan). Code is available at [https://github.com/eugiampetruzzi/abcd_cosinor].

### Cardiac rhythm modeling

A single-component (24-hour) linear mixed-effects cosinor model was fit to minute-level heart rate aggregated to each clock hour across each participant’s valid wear days, yielding three parameters per session: mesor (24-hour rhythm-adjusted mean), amplitude (peak-to-mean swing), and acrophase (clock time of the daily peak; Figure 1). Per-participant best linear unbiased predictions (BLUPs) served as between-person predictors throughout. Within-person stability was indexed by the standard deviation of daily mesor, amplitude, and acrophase among participants with at least seven valid daily fits. The mesor was validated against concurrent cuff-measured and device-computed resting heart rate, and parametric parameters were cross-checked against non-parametric circadian indices (M10, L5, relative amplitude, interdaily stability).

**Figure 1.**
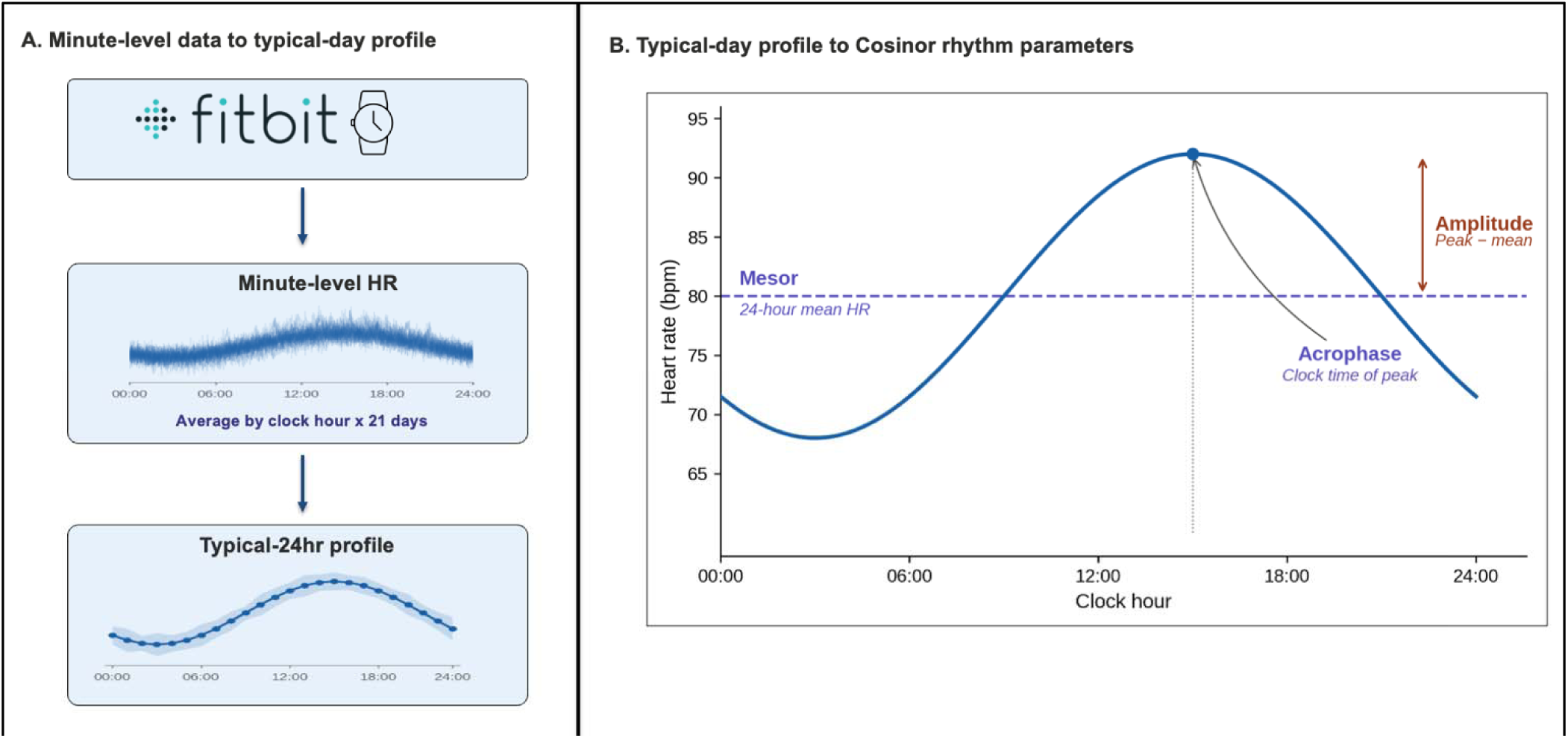
Cosinor modeling of wearable-derived 24-hour heart rate rhythm. ***Note.*** **(A)** Per-participant minute-level heart rate from up to 21 days of Fitbit Charge HR2 wear at Wave 2 (middle) was averaged by clock hour to produce a typical-day profile (bottom). **(B)** A linear mixed-effects cosinor model was fit to the typical-day profile, yielding three rhythm parameters: mesor (24-hour mean), amplitude (peak-to-mean swing), and acrophase (clock time of peak). Per-participant best linear unbiased predicted (BLUP) values for these parameters served as predictors in all subsequent analyses. HR=heart rate; bpm=beats per minute.

### Hypothesis 1: Development of the cardiac rhythm

We estimated age- and sex-specific developmental curves for each parameter using generalized additive mixed models with penalized age splines fit separately by sex, and derived sex-specific centile reference values (GAMLSS). Cross-wave stability was assessed via intraclass correlations and Pearson correlations. To separate intrinsic maturation from behavioral and contextual variation, we refit the age-trajectory models adjusting for physical activity, sleep, pubertal stage (with mediation testing of sex differences), household income, race and ethnicity, cardiac-active medications, and season, reporting the attenuation of the age slope by each factor.

### Hypothesis 2: Prediction of psychopathology and cardiometabolic illness

#### Co-development

To test whether within-person change in cardiac rhythm tracks change in psychopathology and cardiometabolic outcomes, we fit bivariate parallel-process latent growth curve models (lavaan) across Waves 2, 4, and 6, with sex predicting all growth factors, standard errors clustered on site, and full information maximum likelihood for missing data. The key parameter was the slope-slope covariance (whether rhythm parameter changes accompanied greater symptom increase); we also estimated the intercept-intercept covariance and the cross-domain path from Wave 2 rhythm to outcome slope.

### Clinical onset

We fit hierarchical logistic models predicting the first clinical onset of each outcome, defined as crossing the clinical threshold at Wave 4 or Wave 6 while being below threshold at all prior waves; healthy controls (N=1,188) were below threshold at every wave. Covariates were entered in blocks: demographics (M1), baseline subthreshold symptoms (M2), Wave 2 steps and sleep duration (M3), and Wave 2 rhythm (mesor, amplitude, acrophase; M4); within-person instability was entered in parallel M4 models. Standard errors were clustered on family. We report incremental AUC and LRT for the rhythm block (M4 vs. M3) and per-feature odds ratios (per 1-SD) from M4, Benjamini-Hochberg-corrected across 30 tests, and comorbidity among incident cases.

## Results

### Participants

#### Sample characteristics

Demographic and clinical characteristics for the full analytic sample (N=8,301) and the prediction sample (N=2,758) are presented in Table 1. Differences between the two samples are reported in Table S1.

**Table 1.**
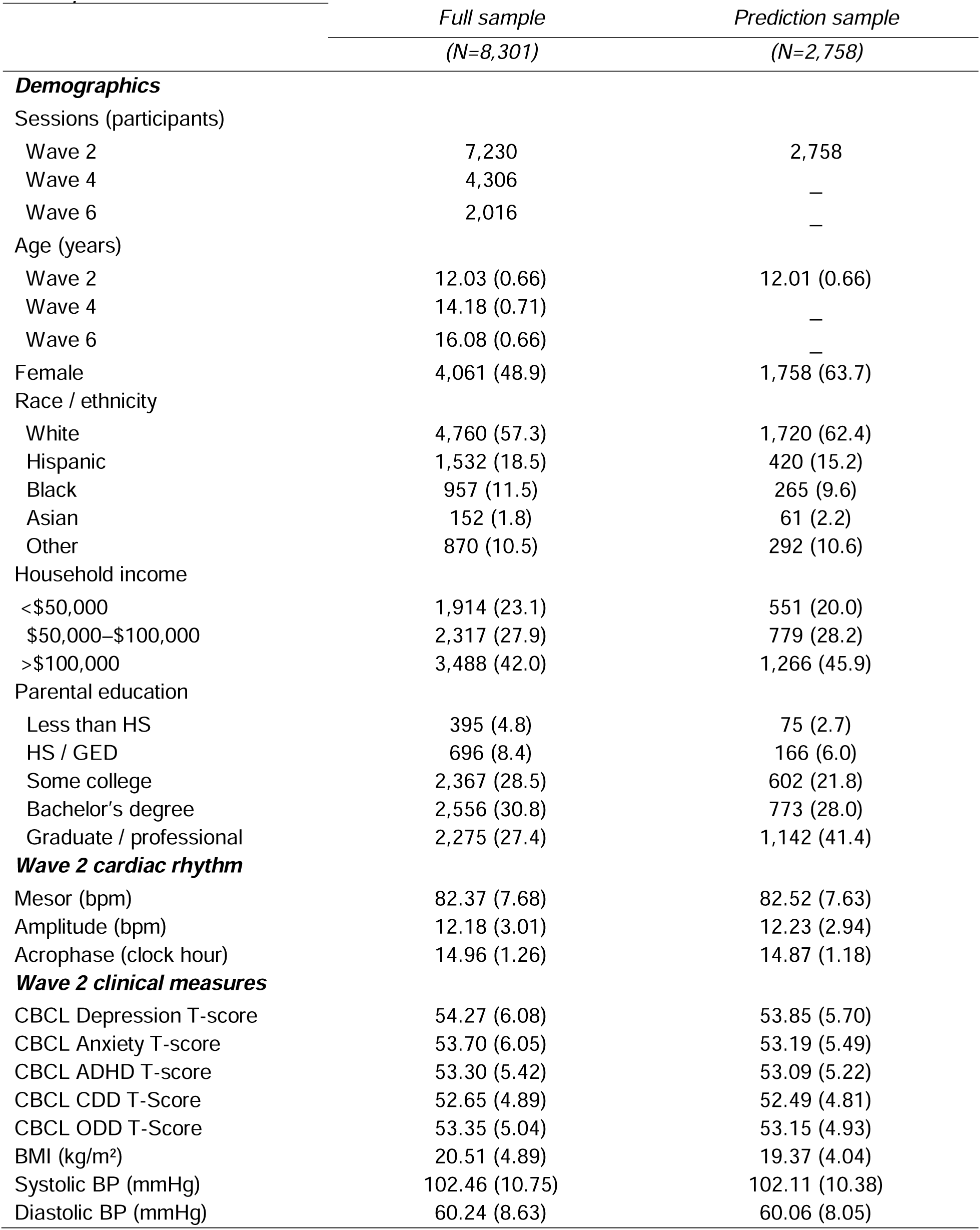
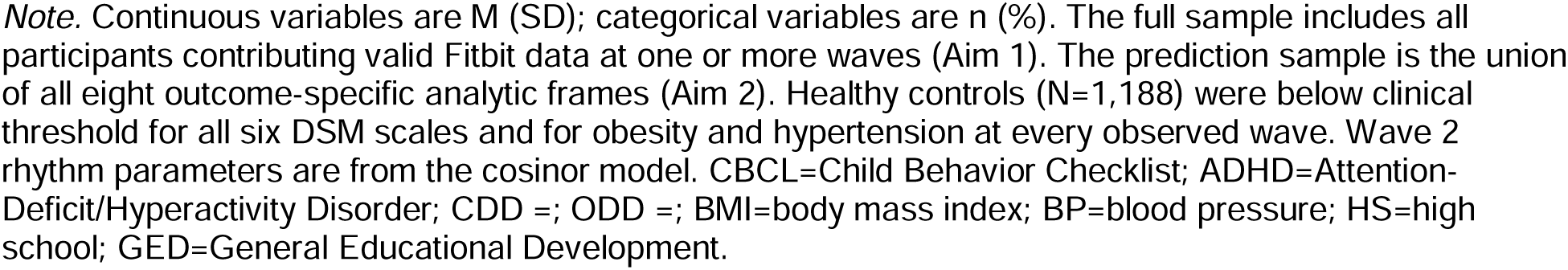
Participant Characteristics.

|  | *Full sample* | *Prediction sample* |
| --- | --- | --- |
|  | *(N=8,301)* | *(N=2,758)* |
| ***Demographics*** |  |  |
| Sessions (participants) |  |  |
| Wave 2 | 7,230 | 2,758 |
| Wave 4 | 4,306 | — |
| Wave 6 | 2,016 | — |
| Age (years) |  |  |
| Wave 2 | 12.03 (0.66) | 12.01 (0.66) |
| Wave 4 | 14.18 (0.71) | — |
| Wave 6 | 16.08 (0.66) | — |
| Female | 4,061 (48.9) | 1,758 (63.7) |
| Race / ethnicity |  |  |
| White | 4,760 (57.3) | 1,720 (62.4) |
| Hispanic | 1,532 (18.5) | 420 (15.2) |
| Black | 957 (11.5) | 265 (9.6) |
| Asian | 152 (1.8) | 61 (2.2) |
| Other | 870 (10.5) | 292 (10.6) |
| Household income |  |  |
| <\$50,000 | 1,914 (23.1) | 551 (20.0) |
| \$50,000–\$100,000 | 2,317 (27.9) | 779 (28.2) |
| >\$100,000 | 3,488 (42.0) | 1,266 (45.9) |
| Parental education |  |  |
| Less than HS | 395 (4.8) | 75 (2.7) |
| HS / GED | 696 (8.4) | 166 (6.0) |
| Some college | 2,367 (28.5) | 602 (21.8) |
| Bachelor's degree | 2,556 (30.8) | 773 (28.0) |
| Graduate / professional | 2,275 (27.4) | 1,142 (41.4) |
| ***Wave 2 cardiac rhythm*** |  |  |
| Mesor (bpm) | 82.37 (7.68) | 82.52 (7.63) |
| Amplitude (bpm) | 12.18 (3.01) | 12.23 (2.94) |
| Acrophase (clock hour) | 14.96 (1.26) | 14.87 (1.18) |
| ***Wave 2 clinical measures*** |  |  |
| CBCL Depression T-score | 54.27 (6.08) | 53.85 (5.70) |
| CBCL Anxiety T-score | 53.70 (6.05) | 53.19 (5.49) |
| CBCL ADHD T-score | 53.30 (5.42) | 53.09 (5.22) |
| CBCL CDD T-Score | 52.65 (4.89) | 52.49 (4.81) |
| CBCL ODD T-Score | 53.35 (5.04) | 53.15 (4.93) |
| BMI (kg/m<sup>2</sup>) | 20.51 (4.89) | 19.37 (4.04) |
| Systolic BP (mmHg) | 102.46 (10.75) | 102.11 (10.38) |
| Diastolic BP (mmHg) | 60.24 (8.63) | 60.06 (8.05) |

*Note.* Continuous variables are M (SD); categorical variables are n (%). The full sample includes all participants contributing valid Fitbit data at one or more waves (Aim 1). The prediction sample is the union of all eight outcome-specific analytic frames (Aim 2). Healthy controls (N=1,188) were below clinical threshold for all six DSM scales and for obesity and hypertension at every observed wave. Wave 2 rhythm parameters are from the cosinor model. CBCL=Child Behavior Checklist; ADHD=Attention-Deficit/Hyperactivity Disorder; CDD =; ODD =; BMI=body mass index; BP=blood pressure; HS=high school; GED=General Educational Development.

### Cardiac rhythm modeling

A single-component (24-hour) cosinor model was fit to 13,552 participant-sessions (8,301 adolescents) across Waves 2 (n=7,230), 4 (n=4,306), and 6 (n=2,016). Fit was excellent (median R²=.85, IQR .78-.89; 0.2% of sessions below the R²=.10 threshold). Mean mesor was 80.3 bpm (SD=8.1), amplitude 11.9 bpm (SD=3.1), and acrophase 15:11 (SD=1.4 h). The mesor correlated strongly with device-computed (r=.93) and concurrent cuff-measured (r=.59) resting heart rate, supporting its validity as an index of cardiac autonomic tone.

### Hypothesis 1: Development of the rhythm

All three rhythm parameters changed significantly across adolescence (Figure 2). Mesor declined −1.80 bpm/year in males and −1.38 bpm/year in females (sex difference p<.001), falling from 83.6 to 72.2 bpm in males and 87.2 to 79.0 bpm in females between ages 10.5 and 18 years; amplitude flattened modestly (−0.21 bpm/year males; −0.16 females) and acrophase shifted later by approximately 10 minutes per year in both sexes. Sex-specific centile reference values are presented in Table 2.

**Figure 2.**
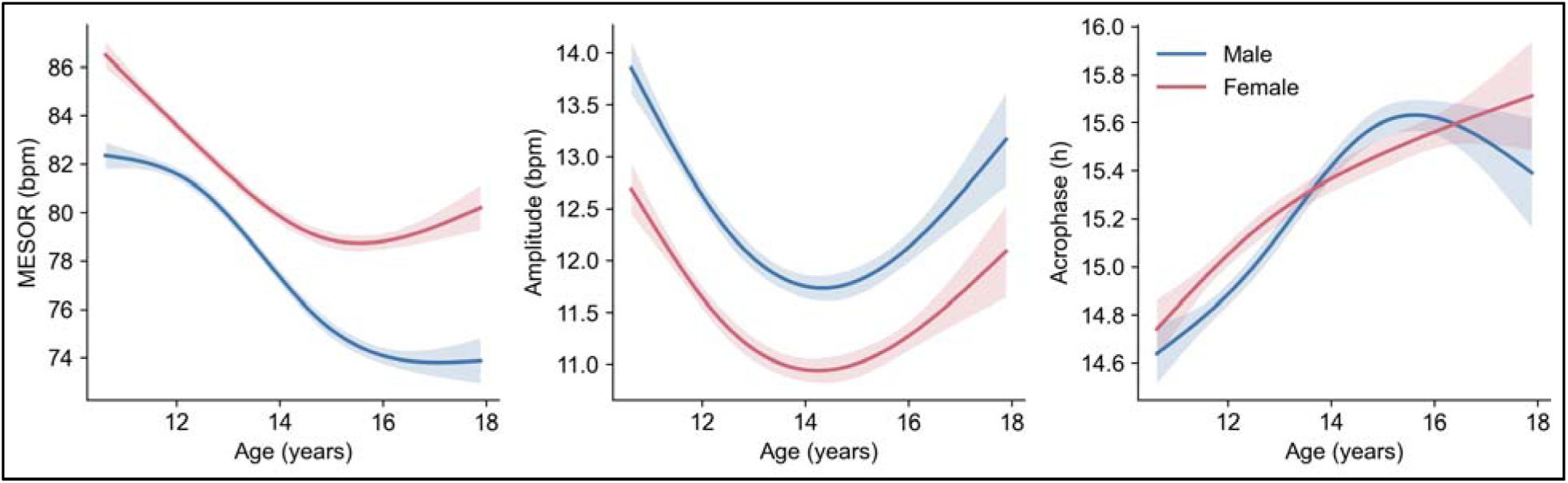
Developmental trajectories of cardiac rhythm parameters by age and sex. *Note.* Developmental curves for mesor (left), amplitude (center), and acrophase (right) estimated from generalized additive mixed models with penalized splines for age, fit separately by sex. Solid lines depict GAMM-predicted means; shaded ribbons depict 95% confidence intervals from cluster-robust standard errors (clustering on ABCD site). Mesor and amplitude decline with age in both sexes, with sex differences widening across adolescence; acrophase shifts later in parallel for males and females. N=8,301 adolescents, 13,552 sessions. bpm=beats per minute; h=hours.

**Table 2.** Sex-specific centile reference values for 24-Hour cardiac rhythm parameter.

| Age<br>(yr) | Sex | 3rd<br>centile | 10th<br>centile | 25th<br>centile | 50th<br>centile | 75th<br>centile | 90th<br>centile | 97th<br>centile |
| --- | --- | --- | --- | --- | --- | --- | --- | --- |
| *Mesor (bpm)* |  |  |  |  |  |  |  |  |
| 10 | Male | 70.3 | 74.9 | 79.5 | 84.4 | 89.2 | 93.3 | 97.3 |
| 12 | Male | 66.9 | 71.5 | 76.2 | 81.4 | 86.5 | 91.2 | 95.7 |
| 14 | Male | 63.3 | 67.6 | 72.1 | 77.3 | 82.6 | 87.6 | 92.7 |
| 16 | Male | 60.7 | 64.5 | 68.7 | 73.6 | 78.9 | 83.9 | 89.3 |
| 18 | Male | 60.1 | 63.6 | 67.5 | 72.2 | 77.5 | 82.8 | 88.5 |
| 10 | Female | 75.2 | 79.4 | 83.7 | 88.4 | 93.1 | 97.4 | 101.6 |
| 12 | Female | 70.0 | 74.3 | 78.7 | 83.6 | 88.4 | 92.8 | 97.2 |
| 14 | Female | 65.7 | 70.2 | 74.7 | 79.8 | 84.8 | 89.3 | 93.8 |
| 16 | Female | 64.1 | 68.7 | 73.4 | 78.6 | 83.8 | 88.5 | 93.1 |
| 18 | Female | 64.1 | 68.9 | 73.7 | 79.0 | 84.4 | 89.2 | 94.0 |
| *Amplitude (bpm)* |  |  |  |  |  |  |  |  |
| 10 | Male | 9.4 | 11.0 | 12.6 | 14.4 | 16.1 | 17.8 | 19.6 |
| 12 | Male | 7.1 | 8.9 | 10.7 | 12.6 | 14.6 | 16.4 | 18.5 |
| 14 | Male | 6.0 | 7.9 | 9.7 | 11.7 | 13.7 | 15.7 | 17.9 |
| 16 | Male | 6.2 | 8.1 | 10.0 | 12.0 | 14.2 | 16.2 | 18.5 |
| 18 | Male | 6.9 | 8.9 | 10.9 | 13.2 | 15.5 | 17.8 | 20.2 |
| 10 | Female | 7.6 | 9.4 | 11.3 | 13.3 | 15.4 | 17.3 | 19.2 |
| 12 | Female | 6.4 | 8.1 | 9.7 | 11.6 | 13.5 | 15.2 | 17.1 |
| 14 | Female | 5.9 | 7.5 | 9.1 | 10.9 | 12.8 | 14.6 | 16.6 |
| 16 | Female | 5.8 | 7.5 | 9.2 | 11.1 | 13.2 | 15.2 | 17.3 |
| 18 | Female | 6.0 | 7.9 | 9.8 | 12.0 | 14.4 | 16.7 | 19.4 |
| *Acrophase (clock hour)* |  |  |  |  |  |  |  |  |
| 10 | Male | 13.18 | 13.62 | 14.05 | 14.57 | 15.19 | 15.92 | 17.03 |
| 12 | Male | 13.02 | 13.56 | 14.09 | 14.71 | 15.44 | 16.32 | 17.61 |
| 14 | Male | 13.25 | 13.94 | 14.60 | 15.35 | 16.21 | 17.23 | 18.68 |
| 16 | Male | 13.17 | 13.95 | 14.68 | 15.47 | 16.34 | 17.30 | 18.61 |
| 18 | Male | 12.89 | 13.83 | 14.64 | 15.49 | 16.38 | 17.33 | 18.56 |
| 10 | Female | 13.06 | 13.46 | 13.91 | 14.49 | 15.19 | 15.99 | 17.06 |
| 12 | Female | 13.16 | 13.67 | 14.21 | 14.88 | 15.66 | 16.54 | 17.68 |
| 14 | Female | 13.09 | 13.77 | 14.45 | 15.24 | 16.15 | 17.19 | 18.59 |
| 16 | Female | 13.20 | 13.89 | 14.56 | 15.34 | 16.23 | 17.24 | 18.65 |
| 18 | Female | 13.62 | 14.25 | 14.88 | 15.65 | 16.58 | 17.69 | 19.34 |

*Note.* Sex-specific centiles of each Wave-2 cardiac rhythm parameter at integer ages, estimated from GAMLSS models with Box-Cox $t$ distributions. Each centile represents the value exceeded by the indicated percentage of same-sex peers at that age (e.g., a male at age 14 with a 75th-centile mesor has a higher 24-h mean heart rate than 75% of same-aged male peers). Mesor and amplitude are in beats per minute; acrophase is clock hour of peak. N=8,301 adolescents, 13,552 sessions. GAMLSS=generalized additive models for location, scale, and shape.

Mesor was the most stable, trait-like parameter across waves (ICC=.56–.61; cross-wave r=.63–.78), followed by amplitude (ICC=.47–.51) and acrophase (ICC=.24–.36). Importantly, individual mesor slopes varied substantially (SD=2.2 bpm/year, exceeding the mean slope), indicating meaningful between-person variation in autonomic development. This change was due largely to intrinsic rather than to behavioral factors: adjusting for daily step count attenuated the amplitude age slope by 40–48% but the mesor slope by only 2–5%, confirming that the decreasing 24-hour mean is not an artifact of declining activity; further, sleep accounted for less than 8% of any parameter. More advanced puberty was independently associated with a lower, flatter, later-peaking rhythm in both sexes (p<.001) and mediated approximately 32% of the amplitude sex difference, consistent with girls being more pubertally advanced at the same chronological age (Shirtcliff et al., 2009). Higher mesor and later acrophase were also observed in Black and lower-income adolescents (p<.001).

### Hypothesis 2: Prediction of psychopathology and cardiometabolic illness

#### Co-development

Within-person change in mesor co-developed most strongly with cardiometabolic outcomes. Mesor and systolic blood pressure trajectories were tightly coupled (slope–slope r=.89, p<.001), as were mesor and BMI trajectories (r=.36, p<.001); BMI was the only outcome for which Wave 2 mesor level prospectively predicted faster outcome growth (cross-domain β=.13, p<.001). Among psychopathology outcomes, mesor change covaried with symptom change for depression (r=.22, p=.022) and ADHD (r=.20, p=.036), but not for anxiety, ODD, or conduct problems. Intercept-intercept covariances were significant for all outcomes (ps<.05). No cross-domain path from mesor level to symptom slope reached significance, indicating that the association of rhythm with psychopathology link is concurrent rather than predictive. See Table S2 for acrophase & amplitude results.

#### Clinical onset

The Wave 2 rhythm block improved prediction of incident onset for all five outcomes beyond demographics, baseline symptoms, and behavior (depression; N=463; ΔAUC=+.020, anxiety; N=310; +.021, externalizing; N=262; +.005, obesity; N=455; +.014, hypertension; N=270; +.013; all LRT p ≤ .01; Table S3). These increments were modest, as expected for a marker added to a strong, autocorrelated symptom baseline (Cook, 2007); the incremental value rests instead on the independent rhythm effects in the fully adjusted model. Higher mesor predicted the first onset of every outcome (depression OR=1.51, anxiety 1.48, externalizing 1.36, obesity 1.47, hypertension 1.54; all p<.01); lower amplitude independently predicted hypertension (0.66) and depression (0.84), and later acrophase predicted anxiety (1.24). Day-to-day instability showed a similar transdiagnostic pattern (Figure 3; Table S4). It is noteworthy that most incident cases were non-comorbid (76.4% met threshold for a single condition; only 9.2% developed both psychopathology and cardiometabolic illness), indicating a shared physiological substrate rather than a small, highly comorbid group.

**Figure 3.**
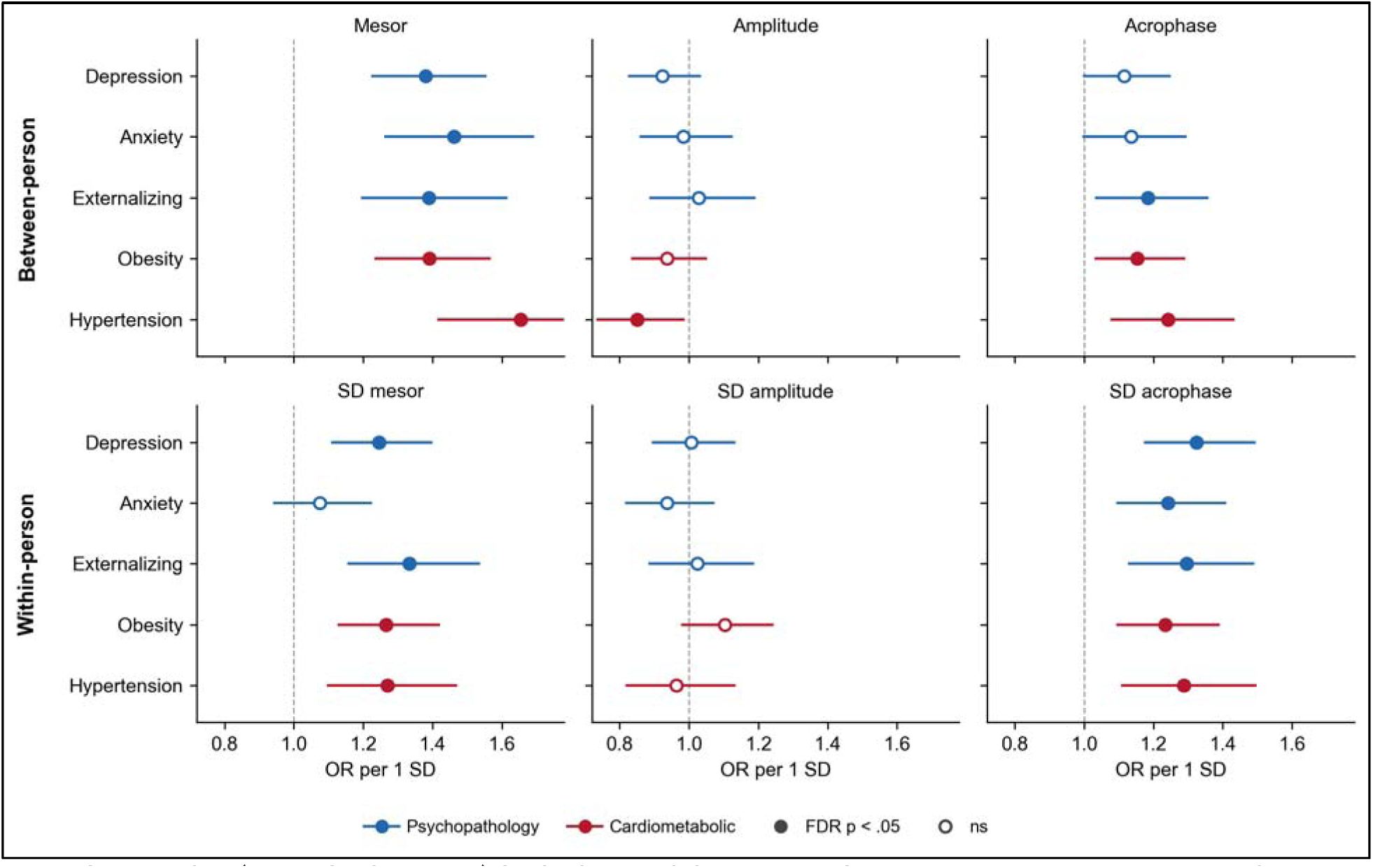
Between- and within-person cardiac rhythm features predicting incident clinical onset across five outcomes. *Note.* Odds ratios (per 1-SD increase) for incident clinical onset of three psychopathology categories (depression, anxiety, combined externalizing) and two cardiometabolic outcomes (obesity, hypertension), relative to the healthy-control pool (n=1,188). The top row shows between-person typical-day cosinor parameters (mesor, amplitude, acrophase); the bottom row shows within-person day-to-day instability (SD of each daily parameter across wear days). Estimates are from logistic regression adjusting for age and sex with family-clustered standard errors. Filled markers indicate Benjamini-Hochberg FDR p<.05 (across 30 tests: 5 outcomes × 6 predictors); open markers are nonsignificant. Horizontal lines are 95% confidence intervals: the dashed line marks OR=1. Blue=psychopathology, red=cardiometabolic. Higher mesor and greater day-to-day acrophase instability predicted onset across nearly all outcomes; amplitude was largely nonpredictive.

As an exploratory convergence check, we repeated the onset analyses using KSADS-COMP diagnoses in place of CBCL clinical elevations for the same three psychopathology categories (Table S5). Replicating the CBCL findings, higher Wave 2 mesor predicted incident diagnostic onset of depression (OR=1.50), anxiety (1.43), and externalizing disorders (1.41; all p<.01); similarly, lower amplitude predicted incident onset of anxiety (OR=0.85) and later acrophase predicted onset of depression (OR=1.18). Findings held with youth-reported diagnoses (Table S6).

## Discussion

We had two goals in this study: first, to characterize the development of the 24-hour cardiac rhythm across adolescence and, second, to test whether individual differences or deviations in this development predict the onset of psychiatric and cardiometabolic illness. Analyzing longitudinal FitBit data from the ABCD Study, we found that the cardiac rhythm changed substantially with age: the 24-hour mean (mesor) declined, the diurnal swing (amplitude) flattened, and the daily peak (acrophase) shifted later. These developmental changes were robust to contextual and behavioral factors, with one important exception: whereas the flattening of amplitude was largely explained by the decline in physical activity over adolescence, the decrease in mesor was almost entirely independent of behavior. Within-person changes in the cardiac rhythm tracked within-person changes in blood pressure, BMI, and symptoms of depression and ADHD. Most notably, the cardiac rhythm in early adolescence predicted the first onset of clinical illness across psychiatric and cardiometabolic domains two to four years later, beyond demographics, baseline symptoms, and behavior. A higher mesor conferred transdiagnostic risk across all five conditions. A flatter, later-peaking cardiac rhythm conferred additional risk, although in an outcome-specific manner: lower amplitude predicted hypertension and depression, and later acrophase predicted anxiety.

These developmental findings extend previous research examining the maturation of cardiac autonomic activity across adolescence. While researchers have characterized the broad course of this maturation (Harteveld et al., 2021; Joyce et al., 2025), the structure of the 24-hour rhythm itself has been described only in cross-sectional pediatric samples (Hadtstein et al., 2004) and small clinical adolescent cohorts (Sigrist et al., 2024). The present study is the first to map the 24-hour rhythm longitudinally and at scale in adolescents, providing developmental growth charts for mesor, amplitude, and acrophase. In addition, individual developmental slopes varied considerably, providing evidence of meaningful variation in adolescent autonomic development that can be observed at the population level. Males showed steeper mesor decline than females, and pubertal development partially mediated this sex difference, indicating that although pubertal stage shapes the developing rhythm, it does not fully account for sex differences in its components (Shirtcliff et al., 2009).

This developmental variation in cardiac rhythm is not merely descriptive, but in addition, indexes transdiagnostic risk for psychiatric and cardiometabolic illness. A higher mesor predicted the first onset of conditions that have typically been examined separately: depression, anxiety, externalizing problems, obesity, and hypertension. These conditions were not generally comorbid in our sample: only 9.2% of incident cases met threshold for both a psychiatric and a cardiometabolic condition. Thus, rather than identifying a comorbid subgroup of adolescents, the rhythm signal appears to be a shared developmental substrate of autonomic imbalance. Importantly, day-to-day instability of the cardiac rhythm, particularly of acrophase, also predicted the first onset of illness across outcomes, indicating that both the level and the stability of the rhythm have transdiagnostic risk. This signal was robust to whether onset was indexed by diagnostic interview or by symptom scale.

We should note two limitations of this study. First, the cardiac rhythm we modeled is a relatively coarse index of autonomic function: it captures the 24-hour structure of heart rate but cannot separate the sympathetic and parasympathetic contributions that shape it because ABCD releases minute-level rather than beat-to-beat data. Incorporating finer-grained cardiac indices — heart rate variability, pre-ejection period, and respiratory sinus arrhythmia — would allow future work to elucidate which autonomic processes are driving the observed associations. Second, ABCD is a community-based cohort and, compared to clinical or high-risk samples, relatively healthy and socioeconomically advantaged. Future research should examine whether the rhythm carries the same prognostic value in adolescents with greater illness and more severe adversity, the populations in whom early detection matters most.

This study also has several strengths. The analytic cohort was large and demographically diverse. Cardiac rhythms were derived from up to 21 days of continuous passive monitoring at each wave rather than from brief laboratory recordings, allowing us to characterize both the typical-day shape and the day-to-day stability of the rhythm across adolescence. The developing 24-hour cardiac rhythm is a passively measurable substrate of adolescent autonomic regulation that, as we documented here for the first time, indexes transdiagnostic risk for the onset of psychiatric and cardiometabolic illness two to four years later. Future research should examine whether continuous passive monitoring across longer developmental windows can identify more precisely the dynamic shifts in autonomic regulation that precede the onset of clinically significant conditions, and whether stratifying samples based on wearable data can lead to earlier, more targeted, and more effective prevention of disorder in youth.

## Data Availability

Access to ABCD Study data is managed through the NIH Brain Development Cohorts Data Use Certification (DUC) process.

https://docs.abcdstudy.org/v/6_0_0/usage/access.html

## Acknowledgments

This research was supported by the National Institute of Mental Health (R37MH101495 to IHG; Intramural Research Project Number ZIA-MH002781 to DSP) and the National Science Foundation Graduate Research Fellowship Program (to EEG). The contributions of the National Institutes of Health (NIH) author(s) are considered Works of the United States Government. The findings and conclusions presented in this paper are those of the author(s) and do not necessarily reflect the views of the NIH or the U.S. Department of Health and Human Services.

