## Supplement for "Cardiac rhythm development: A wearable device index of risk for physical and mental illness in adolescence"

**Table S1**

*Comparison of the Prediction Sample and the Remainder of the Full Cosinor Cohort*

| Characteristic | Prediction sample (n = 2,758) | Remainder (n = 4,472) | p |
| --- | --- | --- | --- |
| Age, Wave 2 (years) | 12.00 (0.66) | 12.03 (0.66) | 0.134 |
| Female | 1,758 (63.7) | 1,785 (39.9) | <.001 |
| Race / ethnicity |  |  |  |
| White | 1,720 (62.4) | 2,543 (56.9) | <.001 |
| Hispanic | 420 (15.2) | 901 (20.2) |  |
| Black | 265 (9.6) | 517 (11.6) |  |
| Asian | 61 (2.2) | 71 (1.6) |  |
| Other | 292 (10.6) | 439 (9.8) |  |
| Household income |  |  |  |
| < $50,000 | 551 (20.0) | 1,051 (23.5) | <.001 |
| $50,000–$100,000 | 779 (28.2) | 1,268 (28.4) |  |
| > $100,000 | 1,266 (45.9) | 1,828 (40.9) |  |
| Parental education |  |  |  |
| Less than HS | 75 (2.7) | 155 (3.5) | <.001 |
| HS / GED | 166 (6.0) | 333 (7.4) |  |
| Some college | 602 (21.8) | 1,143 (25.6) |  |
| Bachelor's degree | 773 (28.0) | 1,260 (28.2) |  |
| Graduate / professional | 1,142 (41.4) | 1,581 (35.4) |  |
| Wave 2 mesor (bpm) | 82.52 (7.63) | 82.29 (7.71) | 0.217 |
| Wave 2 amplitude (bpm) | 12.23 (2.94) | 12.14 (3.05) | 0.226 |
| Wave 2 acrophase (clock hour) | 14.87 (1.18) | 15.01 (1.30) | <.001 |
| CBCL Depression T | 53.85 (5.70) | 54.55 (6.32) | <.001 |
| CBCL Anxiety T | 53.19 (5.49) | 54.04 (6.38) | <.001 |
| CBCL ADHD T | 53.09 (5.22) | 53.44 (5.54) | 0.008 |
| BMI (kg/m²) | 19.37 (4.04) | 21.23 (5.23) | <.001 |
| Systolic BP (mmHg) | 102.11 (10.38) | 102.67 (10.98) | 0.134 |
| Diastolic BP (mmHg) | 60.06 (8.05) | 60.36 (8.98) | 0.325 |

*Note.* Continuous variables are M (SD); categorical variables are n (%). The prediction sample is the union of the five outcome-specific onset frames (Aim 2); the remainder comprises Wave-2 cosinor-cohort members not entering any onset analysis. p values are from Welch t tests (continuous) and chi-square tests (categorical; test reported on the first level of each block). CBCL = Child Behavior Checklist; ADHD = Attention-Deficit/Hyperactivity Disorder; BMI = body mass index; BP = blood pressure.

**Table S2**

*Co-Development of Mesor and Health-Outcome Trajectories: Bivariate Parallel-Process Latent Growth Models*

| Outcome | Slope-slope r | Intercept-intercept r | Cross-domain β |
| --- | --- | --- | --- |
| Systolic blood pressure | 0.89*** | 0.07* | -0.10 |
| Body mass index | 0.36*** | 0.21*** | +0.13*** |
| Depression | 0.22* | 0.13*** | +0.00 |
| ADHD | 0.20* | 0.15*** | -0.00 |
| Anxiety | 0.07 | 0.07*** | +0.03 |
| ODD | 0.05 | 0.10*** | -0.02 |
| Conduct | -0.00 | 0.13*** | -0.01 |

*Note.* Bivariate parallel-process latent growth models estimating mesor and outcome trajectories jointly across Waves 2, 4, and 6 (MLR estimator, full-information maximum likelihood, site-clustered SEs), with sex predicting all growth factors. Slope-slope r = standardized covariance between mesor slope and outcome slope (positive = slower mesor decline tracks greater outcome increase); intercept-intercept r = concurrent level association; cross-domain β = Wave-2 mesor level predicting outcome slope. Outcomes are ordered by slope-slope effect size. ADHD = Attention-Deficit/Hyperactivity Disorder; ODD = Oppositional Defiant Disorder. * p < .05. ** p < .01. *** p < .001.

**Table S3**

*Incremental Prediction of Clinical Onset by Wave-2 Cardiac Rhythm (Hierarchical M1–M4)*

| Outcome | Cases/N | AUC M3 | AUC M4 | ΔAUC | LRT p | Mesor | Amplitude | Acrophase |
| --- | --- | --- | --- | --- | --- | --- | --- | --- |
| Depression | 322/1135 | 0.744 | 0.764 | +0.020 | 1.69e-05*** | 1.51 [1.27, 1.80]*** | 0.84 [0.71, 0.99]* | 1.02 [0.87, 1.19] |
| Anxiety | 221/943 | 0.755 | 0.776 | +0.021 | 2.19e-05*** | 1.48 [1.21, 1.82]*** | 0.83 [0.68, 1.01] | 1.24 [1.04, 1.49]* |
| Externalizing | 171/954 | 0.825 | 0.831 | +0.005 | 0.0127* | 1.36 [1.08, 1.70]** | 0.84 [0.66, 1.06] | 1.14 [0.94, 1.38] |
| Obesity | 312/1123 | 0.767 | 0.781 | +0.014 | 9.98e-05*** | 1.47 [1.23, 1.75]*** | 0.92 [0.78, 1.08] | 1.07 [0.92, 1.25] |
| Hypertension | 160/453 | 0.846 | 0.859 | +0.013 | 0.000424*** | 1.54 [1.18, 2.00]** | 0.66 [0.49, 0.89]** | 1.24 [0.94, 1.62] |

*Note.* Rhythm cells are odds ratios [95% CI] per 1-SD from the fully adjusted model (M4). Hierarchical logistic models versus the healthy-control pool (n = 1,188), family-clustered SEs. Blocks: M1 demographics; M2 + baseline symptoms (Wave-0 T-score, or Wave-0 BMI / Wave-2 systolic BP for cardiometabolic outcomes); M3 + behavior (Wave-2 sleep, MVPA); M4 + rhythm. ΔAUC and the likelihood-ratio test (LRT) compare M4 with M3. Externalizing = onset on any of ADHD, ODD, or Conduct. AUC = area under the receiver operating characteristic curve. * p < .05. ** p < .01. *** p < .001.

**Table S4**

*Transdiagnostic Specificity: Between- and Within-Person Rhythm Features Predicting Incident Onset*

| Outcome | Mesor | Amplitude | Acrophase |
| --- | --- | --- | --- |
| *Between-person* |  |  |  |
| Depression | 1.38 [1.23, 1.55]*** | 0.92 [0.83, 1.03] | 1.12 [1.00, 1.25] |
| Anxiety | 1.46 [1.26, 1.69]*** | 0.98 [0.86, 1.12] | 1.14 [1.00, 1.29] |
| Externalizing | 1.39 [1.20, 1.61]*** | 1.03 [0.89, 1.19] | 1.18 [1.03, 1.36]* |
| Obesity | 1.39 [1.24, 1.56]*** | 0.94 [0.84, 1.05] | 1.15 [1.03, 1.29]* |
| Hypertension | 1.65 [1.42, 1.93]*** | 0.85 [0.74, 0.98]* | 1.24 [1.08, 1.43]** |
| *Within-person* |  |  |  |
| Depression | 1.25 [1.11, 1.40]*** | 1.01 [0.90, 1.13] | 1.32 [1.18, 1.49]*** |
| Anxiety | 1.07 [0.94, 1.22] | 0.94 [0.82, 1.07] | 1.24 [1.10, 1.41]** |
| Externalizing | 1.33 [1.16, 1.53]*** | 1.02 [0.89, 1.18] | 1.30 [1.13, 1.49]*** |
| Obesity | 1.27 [1.13, 1.42]*** | 1.10 [0.98, 1.24] | 1.23 [1.10, 1.39]** |
| Hypertension | 1.27 [1.10, 1.47]** | 0.96 [0.82, 1.13] | 1.29 [1.11, 1.49]** |

*Note.* Cells are odds ratios [95% CI] per 1-SD. Logistic models versus the healthy-control pool (n = 1,188), adjusting for age and sex with family-clustered SEs. Between-person predictors are the typical-day cosinor parameters; within-person predictors are the standard deviations of daily mesor, amplitude, and acrophase among participants with 7+ valid daily fits. Significance reflects Benjamini-Hochberg FDR correction across all 30 tests (5 outcomes × 6 predictors). Estimates are those summarized in Figure 3. * p < .05. ** p < .01. *** p < .001.

**Table S5**

*KSADS-COMP Diagnostic Onset Predicted by Wave-2 Cardiac Rhythm (Exploratory Convergent Check)*

| Category | Cases | Mesor | Amplitude | Acrophase |
| --- | --- | --- | --- | --- |
| *CBCL healthy controls (n = 1,188)* |  |  |  |  |
| Depression | 561 | 1.50 [1.34, 1.69]*** | 0.89 [0.79, 1.00] | 1.18 [1.04, 1.34]** |
| Anxiety | 414 | 1.43 [1.25, 1.63]*** | 0.85 [0.75, 0.97]* | 1.00 [0.88, 1.15] |
| Externalizing | 210 | 1.41 [1.19, 1.68]*** | 1.06 [0.90, 1.24] | 1.05 [0.87, 1.26] |
| *KSADS healthy controls (n = 2,587)* |  |  |  |  |
| Depression | 561 | 1.24 [1.12, 1.37]*** | 1.04 [0.94, 1.15] | 1.02 [0.93, 1.13] |
| Anxiety | 414 | 1.15 [1.03, 1.28]* | 0.99 [0.88, 1.11] | 0.91 [0.82, 1.01] |
| Externalizing | 210 | 1.18 [1.02, 1.37]* | 1.22 [1.05, 1.42]* | 0.94 [0.82, 1.08] |

*Note.* Cells are odds ratios [95% CI] per 1-SD from logistic models adjusting for age and sex with family-clustered SEs (lenient case definition). KSADS onset = first met criteria (present, past, or partial remission) on the parent KSADS-COMP at Year 4 or Year 6 with documented absence at Baseline and Year 2. Anxiety = generalized anxiety, separation anxiety, social anxiety, panic, and agoraphobia; Externalizing = ADHD, oppositional defiant, or conduct disorder. The two panels differ only in the control pool. KSADS = Kiddie Schedule for Affective Disorders and Schizophrenia. * p < .05. ** p < .01. *** p < .001.

**Table S6**

*Child-Report KSADS-COMP Diagnostic Onset Predicted by Wave-2 Cardiac Rhythm (Exploratory Convergent Check)*

| Category | Cases | Mesor | Amplitude | Acrophase |
| --- | --- | --- | --- | --- |
| *CBCL healthy controls (n = 1,188)* |  |  |  |  |
| Depression | 1199 | 1.42 [1.29, 1.57]*** | 0.80 [0.73, 0.89]*** | 1.25 [1.13, 1.38]*** |
| Anxiety | 851 | 1.40 [1.27, 1.55]*** | 0.80 [0.73, 0.89]*** | 1.09 [0.99, 1.21] |
| Externalizing | 310 | 1.46 [1.25, 1.69]*** | 0.99 [0.84, 1.15] | 1.42 [1.21, 1.66]*** |
| *Youth-KSADS healthy controls (n = 3,464)* |  |  |  |  |
| Depression | 1199 | 1.13 [1.05, 1.21]** | 0.93 [0.86, 1.00]* | 1.10 [1.03, 1.19]** |
| Anxiety | 851 | 1.12 [1.03, 1.21]** | 0.90 [0.83, 0.98]* | 0.97 [0.89, 1.06] |
| Externalizing | 310 | 1.18 [1.05, 1.33]** | 1.03 [0.92, 1.17] | 1.24 [1.11, 1.40]*** |

*Note.* Cells are odds ratios [95% CI] per 1-SD; parallel to Table S5 but using the youth self-report KSADS-COMP (lenient case definition; age and sex adjusted, family-clustered SEs). The youth interview covers fewer modules: Anxiety = generalized anxiety, social anxiety, and panic; Externalizing = conduct disorder only (youth report has no ADHD or oppositional defiant disorder, and conduct is not assessed before Year 4, so the conservative definition is used). Youth self-report endorses threshold more often than parent report, so case counts are not comparable to Table S5. The two panels differ only in the control pool. * p < .05. ** p < .01. *** p < .001.
